# Immunogenicity of Pfizer-BioNTech COVID-19 mRNA Primary Vaccination Series in Recovered Individuals Depends on Symptoms at Initial Infection

**DOI:** 10.1101/2022.03.29.22272714

**Authors:** Sabryna Nantel, Benoîte Bourdin, Kelsey Adams, Julie Carbonneau, Henintsoa Rabezanahary, Marie-Ève Hamelin, Deirdre McCormack, Patrice Savard, Yves Longtin, Matthew P. Cheng, Gaston De Serres, Jacques Corbeil, Vladimir Gilca, Mariana Baz, Guy Boivin, Caroline Quach, Hélène Decaluwe

## Abstract

**Importance:** Public health vaccination recommendations for COVID-19 primary series and boosters in previously infected individuals differ worldwide. As infection with SARS-CoV-2 is often asymptomatic, it remains to be determined if vaccine immunogenicity is comparable in all previously infected subjects. We present detailed immunological evidence to clarify the requirements for one-or two-dose primary vaccination series for naturally primed individuals.

**Objective:** Evaluate the immune response to COVID-19 mRNA vaccines in healthcare workers (HCWs) who recovered from a SARS-CoV-2 infection.

**Design:** Multicentric observational prospective cohort study of HCWs with a PCR-confirmed SARS-CoV-2 infection designed to evaluate the dynamics of T and B cells immune responses to primary infection and COVID-19 mRNA vaccination over 12 months.

**Participants:** Unvaccinated HCWs with PCR-confirmed SARS-CoV-2 infection were selected based on the presence or absence of symptoms at infection and serostatus at enrollment. Age- and sex-matched adults not infected with SARS-CoV-2 prior to vaccination were included as naïve controls.

**Exposure:** Vaccination with Pfizer BioNTech BNT162b2 mRNA vaccine.

**Main Outcome(s) and Measure(s):** Immunity score (zero to three), before and after vaccination, based on anti-RBD IgG ratio, serum capacity to neutralize live virus and IFN-γ secretion capacity in response to SARS-CoV-2 peptide pools above the positivity threshold for each of the three assays. We compared the immunity score between groups based on subjects’ symptoms at diagnosis and/or serostatus prior to vaccination.

**Results:** None of the naïve participants (n=14) showed a maximal immunity score of three following one dose of vaccine compared to 84% of the previously infected participants (n=55). All recovered individuals who did not have an immunity score of three were seronegative prior to vaccination, and 67% had not reported symptoms resulting from their initial infection. Following one dose of vaccine, their immune responses were comparable to naïve individuals, with significantly weaker responses than those who were symptomatic during infection.

**Conclusions and Relevance:** Individuals who did not develop symptoms during their initial SARS-CoV-2 infection and were seronegative prior to vaccination present immune responses comparable to that of naïve individuals. These findings highlight the importance of administering the complete two-dose primary regimen and following boosters of mRNA vaccines to individuals who experienced asymptomatic SARS-CoV-2 infection.

**KEY POINTS:** 

**Question:** Is a single dose of COVID-19 mRNA vaccine sufficient to induce robust immune responses in individuals with prior SARS-CoV-2 infection?

**Findings:** In this cohort of 55 health care workers previously infected with SARS-CoV-2, we show that the absence of symptoms during initial infection and negative serostatus prior to vaccination predict the strength of immune responses to COVID-19 mRNA vaccine. Lack of symptoms and a negative serostatus prior to vaccination leads to immune responses comparable to naïve individuals.

**Meaning:** Our results support a two-dose primary series requirement for any individual with prior history of asymptomatic SARS-CoV-2 infection.

## INTRODUCTION

As of March 2022, more than 450 million people have been infected by the Severe Acute Respiratory Syndrome Coronavirus 2 (SARS-CoV-2), leading to over six million deaths from Coronavirus Infectious Disease 2019 (COVID-19).^1-3^ The range of symptoms associated with SARS-CoV-2 infection is highly diverse, and the scale of symptoms spans from asymptomatic to severe.^4^ Public health vaccination guidelines of adults who have recovered from SARS-CoV-2 infection differ depending on the country. In Canada and the United States, the National Advisory Committee on Immunization (NACI) and the Centers for Disease Control and Prevention (CDC) recommend a two-dose primary regimen for every adult,^5-8^ regardless of previous infection status, while countries such as France, Germany, Italy, and Switzerland consider that infection with SARS-CoV-2 is equivalent to one vaccine dose.^9^ It remains to be established if vaccine requirements for previously infected subjects differ from those for naïve individuals.

Up to one year after their infection, recovered individuals present strong and slowly decaying humoral and cellular responses.^10-13^ Importantly, it was suggested that the strength of the immune response following natural infection is proportional to the disease severity.^14-16^ In previously infected individuals, one vaccine dose a few months after infection acts as a booster of immune responses.^17,18^ It elicits much stronger B- and T-cell-specific responses in previously infected individuals, compared to naïve individuals, with minimal benefits observed when a second dose is given 21 days after the first one.^10,11,18-22^ However, it is unknown if individuals who were previously infected, but did not experience symptoms, show similar heightened immune responses after a single mRNA vaccine dose. We hypothesized that the symptom severity during natural infection impacts the response to vaccination and that individuals who did not experience symptoms during infection may present suboptimal protection after one dose of vaccine.

In this report, we used three biomarkers of immunity to characterize an effective immune response to SARS-CoV-2. We assessed (1) circulating anti-RBD IgG levels, (2) functional serum capacity to neutralize live SARS-CoV-2 virus and (3) IFN-γ secretion by memory cells in response to mega pools of SARS-CoV-2 peptides in a cohort of naïve and previously infected individuals before and after COVID-19 mRNA vaccination. We compared the impact of symptoms at the time of primary infection and serology at enrollment (prior to vaccination) on the immune responses to one and two vaccine doses.

## METHODS

### Study Participants

Our cohort consisted of 55 previously infected health care workers (HCWs) who were recruited following a PCR-confirmed SARS-CoV-2 infection as part of the RECOVER study (n=569),^23^ and 14 naïve HCWs with neither history of SARS-CoV-2 infection nor the presence of anti-N or anti-RBD antibodies against SARS-CoV-2 prior to vaccination with BNT162b2 mRNA vaccine (Pfizer-BioNTech, 30 μg per dose). Participants were selected based on their symptomatology at the time of infection and their serostatus at enrollment in the study. All RECOVER subjects who were asymptomatic and for whom samples were available before and after vaccination were included in the study (n= 19). Participants who were symptomatic during infection (n=26) were randomly selected from the rest of the RECOVER cohort based on their serostatus at enrollment after being matched with participants from the asymptomatic group for sex, age, time since infection and time between infection and vaccination. Characteristics of the study population are shown in (**eTable1 in Supplemental 1)**. The severity of the initial SARS-CoV-2 infection was determined using the WHO clinical progression scale.^4^ Blood samples for humoral and cellular immunity were collected at enrollment and at different time points after vaccination as described in **eFigure 1 in Supplemental 1**. Participants were recruited from August 17, 2020, to April 8, 2021, and followed for one year after enrollment.

### Sample Collection and Processing

Blood samples were collected into serum separation tubes (SST(tm), BD) and acid–citrate–dextrose tubes (ACD, BD). They were shipped to the Rare Pediatric Disease (RaPID) biobank at the CHU Sainte-Justine, where serum and peripheral blood mononuclear cells (PBMCs) were isolated according to standard operation procedures (SOPs) using SepMate(tm) tubes (Stemcell Technologies, Canada) for PBMCs isolation. Serum was cryopreserved at -80°C, and PBMCs were cryopreserved in complete RPMI (Gibco) with 10% DMSO and stored in liquid nitrogen until used.

### Exposures and Outcome

Exposure was defined as vaccination with Pfizer BioNTech BNT162b2 mRNA vaccine. Vaccine administration was not part of the study itself, as participants received their vaccines through routine public health programs. The primary outcome was the plurality of immune responses developed after each vaccine dose, represented as an immunity score from zero to three. One point was attributed for each of the following immune assays above positive threshold : anti-RBD IgG levels by ELISA, plasma capacity to neutralize live SARS-CoV-2 (ancestral strain) by micro-neutralization assay, and IFN-γ secretion by PBMCs in response to SARS-CoV-2 mega pool of Spike peptides, determined by ELISpot.

### Enzyme-Linked ImmunoSorbent Assay (ELISA)

An ELISA was performed on serum samples to detect specific IgG antibodies for the Receptor Binding Domain (anti-RBD) of SARS-CoV-2 Spike Glycoprotein as previously described.^24,25^ Seropositivity was defined as an OD_490_ ratio greater than one with the experimentally determined cut-off, which corresponds to the mean of negative controls plus three times the standard deviation of the negative controls. This anti-RBD IgG ELISA was validated on pre-pandemic samples (negative controls) and samples from subjects with confirmed SARS-CoV-2 infection sent by the Canadian National Microbiology Laboratory.^23^ Absence of previous infection to SARS-CoV-2 was confirmed in the naïve cohort by performing an ELISA detecting specific IgG antibodies for the Nucleoprotein (anti-N) of SARS-CoV-2 and the anti-RBD ELISA before vaccination.

### Micro-Neutralization Assay

Neutralizing antibody titers were assessed using SARS-CoV-2/Québec City/21697/2020 strain (ancestral Wuhan-1 like SARS-CoV-2), isolated from a clinical sample in March 2020 in Québec City, Canada. Two-fold dilutions of heat-inactivated serum were prepared, starting with 1:20 dilution. Equal volumes of serum and virus were mixed and incubated for one hour at room temperature. The residual infectivity of those mixtures was assessed in quadruplicate wells of African green monkey kidney E6 cell line (Vero ATCC® CRL-1586™). Neutralizing antibody titers were defined as the reciprocal of the serum dilution that completely neutralized the infectivity of 100 TCID_50_ of SARS-CoV-2, determined by the absence of cytopathic effect on cells after four days.^26^ The neutralizing antibody titer is calculated using the Reed/Muench method.^27^ These studies were performed in the Containment Level 3 (CL3) laboratory at the CHU de Québec-Université Laval.

### Enzyme-Linked ImmunoSpot Assay (ELISpot)

Cell-mediated immune (CMI) response was estimated by ELISpot. Frozen PBMCs were rapidly thawed and rested overnight. They were then stimulated with 1 µg/mL of spike glycoprotein (S), nucleocapsid protein (NCAP) or membrane protein (VME1) mega pools of SARS-CoV-2 peptides from the ancestral Wuhan-1 like strain (JPT Peptide Technologies, JPT) as described elsewhere.^28^ Spots were revealed using BIO-RAD Alkaline Phosphatase Conjugate Substrate Kit. The resulting ELISpots were analyzed using CTL ImmunoSpot® S5 UV Analyzer (Cellular Technology Ltd, OH). Culture media and phytohemagglutinin (PHA, Sigma) were used as negative and positive controls, respectively. The positive threshold response was defined as more than 25 spot-forming units (SFU) per million cells.

### Activation Induced Markers (AIM) Assay

Thawed and rested PBMCs were stimulated for 20 hours with 1 µg/mL of the CD4-Spike peptides mega pool developed and synthesized by the Sette laboratory.^29,30^ Constituted of 246 HLA-class II restricted 15-mers spanning the whole Spike Glycoprotein, the CD4-Spike mega pool stimulates both CD4 and CD8 T cells. Cells were then stained with panels of fluorescent monoclonal antibodies (**eTable 3 in Supplemental 1**). Data were acquired with an LSR FORTESSA II with High Throughput Sampler (HTS) from BD Biosciences (**eFigure 2 in Supplemental 1** for gating strategy). FlowJo software, version 10.7.1, was used to perform all data analysis. Fluorescence Minus One (FMO), unstimulated (negative control) and PMA-Ionomycine (positive control) conditions were used to set the gates for each participant. Results are represented by the frequency of activated cells following peptide stimulation minus the frequency of activated cells in the unstimulated condition.

### Statistics

All statistical tests were performed using Prism 9, version 9.2.0 (2021 GraphPad Software, LLC) and data were displayed as mean ± SEM. Wilcoxon-Mann-Whitney or Kruskal-Wallis tests were used to evaluate statistical significance between groups for variables that did not follow a normal distribution. Simple linear regression is presented with Pearson correlation (R^2^) and P-value (p). Significance was set as *p <.05; **p <.01; ***p <.001; ****p <.0001.

### Ethics

RECOVER protocols were approved by the Research Ethics Board (REB) at the CHU Sainte-Justine (CHUSJ) under study MP-21-2021-3035 and in each of the five participating centers in the Province of Québec. Written informed consent was obtained from all participants during the recruitment period, and ongoing consent was reviewed at each subsequent visit.

## RESULTS

### Participants and immune characteristics after initial infection

The RECOVER cohort consists of 569 HCWs infected with SARS-CoV-2 during the first and second pandemic waves.^23^ Overall, 95% of participants had symptoms at the time of diagnosis (n=541), and very few were asymptomatic (n=28). Among asymptomatic subjects, only 43% were seropositive at enrollment, in contrast to 77% of symptomatic individuals (**eFigure 3A in Supplemental 1**). The presence of symptoms at the time of initial infection correlated with more robust anti-RBD IgG levels (mean ± SEM, 2.8±0.4 vs. 1.1±0.1, P<.001) in contrast to subjects who did not experience symptoms. Further, antecedent infection led to long-lasting humoral immune responses in most HCWs (up to 12 months post-infection; **eFigure 3B in Supplemental 1**). Disease severity, assessed from 1 to 5 using the WHO clinical progression score,^4^ increased the strength of this humoral response in HCWs (**eFigure 3C in Supplemental 1**).

Cell-mediated immune responses to the major components of SARS-CoV-2 were also assessed in almost half of the RECOVER cohort (n=200) by ELISpot. Responses to Spike, NCAP and VME1 were present and maintained up to one year after natural infection in the majority of participants (84%, 80% and 72% respectively), with the most potent response being specific for Spike antigens (140.3 ± 12.7 SFU/10^6^ PBMCs, compared to 86.17 ± 6.7 and 95.04 ± 9.4 SFU/10^6^ PBMCs for NCAP and VME1) (**eFigure 4 in Supplemental 1**).

We selected a subset of individuals (n=55) to study the impact of previous infection on the immune response to vaccination. All subjects received the BNT162b2 Pfizer BioNTech COVID-19 vaccine, with a first dose administered 8.4 ± 2.3 months (range: 0.8-13.1 months) after infection (**eTable2 in Supplemental 1**). The second dose was offered 16 ± 6 weeks after the first dose. Samples were collected before vaccination, around 45 ± 25 days and 57 ± 30 days after the first and second dose, respectively. They were compared to 14 previously naïve individuals before vaccination, 29 ± 2 days after the first vaccine dose and 35 ± 5 days after the second dose.

### One vaccine dose strongly boosts humoral and cellular immune responses in recovered individuals

Prior to vaccination, most recovered HCWs presented detectable anti-RBD IgG levels (1.6 ± 0.2) and IFN-γ responses to Spike antigens (135.9 ± 30.1 SFU/10^6^ PBMCs), while serum neutralization capability was mainly absent or low. Following one vaccine dose, recovered individuals demonstrated a striking increase in their anti-RBD IgG levels, neutralization capacity and cell-mediated immune response (1.6 ± 0.2 to 6.3 ± 0.3, P<.001, 17.2 ± 1.3 to 502.9 ± 91.3, P<.001, 135.9 ± 30.1 to 278.5 ± 40.7 SFU/10^6^ PBMCs, P<.001) (**Figure 1A)**. In contrast, only one naïve individual showed detectable neutralization titers following the first vaccine dose. In naïve subjects, the breadth of humoral and cellular immune response was also significantly reduced compared to previously infected subjects, with reduced IgG levels (2.0 ± 0.2) and a lower number of IFN-γ secreting cells (68.2 ± 13.6 SFU/10^6^ PBMCs) compared to recovered HCWs (6.3 ± 0.3 IgG OD ratio and 278.5 ± 40.7 SFU/10^6^ PBMCs respectively; P<.001 and P=.03) (**eFigure 5 in Supplemental 1**).

**Figure 1:**
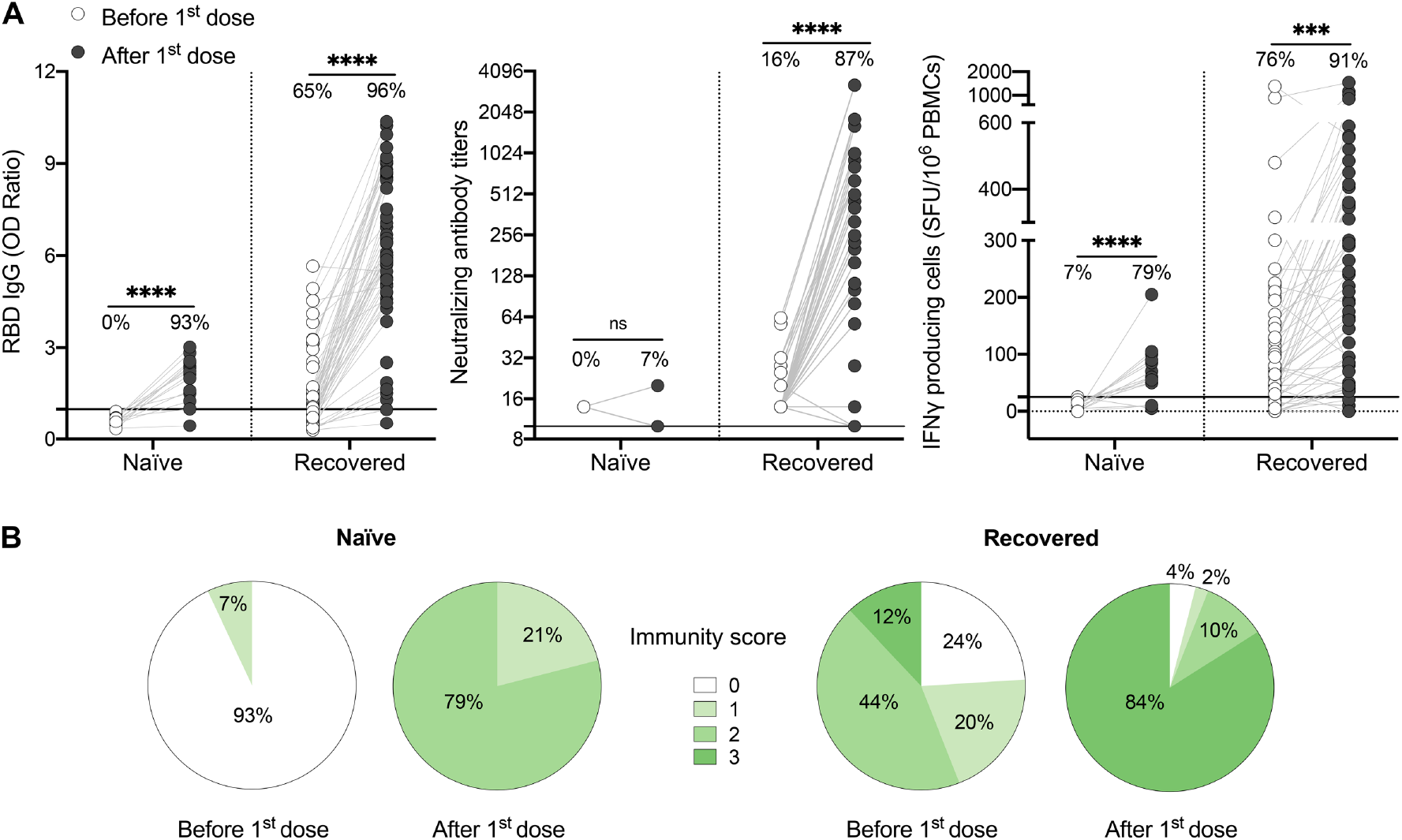
One dose of COVID-19 mRNA-vaccine strongly boosts SARS-CoV-2-specific humoral and cellular responses in most HCWs who recovered from SARS-CoV-2 infection. (**A**) SARS-CoV-2 Spike RBD–specific binding IgG levels assessed by ELISA (left panel), SARS-CoV-2 neutralizing antibody titers (middle panel), IFN-γ secreting cells per million in response to SARS-CoV-2 Spike glycoprotein peptides (right panel) assessed by ELISpot for naïve (n=14) and recovered HCWs (n=55) before (white circles) and after one dose of vaccine (black circles). The black line indicates the positive threshold value, and the percentage of individuals with responses above positive threshold value is indicated for each time point. Statistical significance was assessed by Wilcoxon-Mann-Whitney tests. *P <.05; **P <.01; ***P <.001; ****P <.0001. (**B**) Pie charts of calculated immunity score for naïve (n=14) and recovered HCWs (n=55) before and after one dose of vaccine.

It has been demonstrated that a well-coordinated humoral and cellular immune response, with the development of both antibody neutralizing capacity and cell-mediated immune responses, protects against severe disease.^14,15^ In previously infected HCWs vaccinated with one dose, we observed that the strength of the cellular response, as assessed by ELISpot assay, correlates positively with both RBD-binding IgG and SARS-CoV-2 neutralizing antibody titers (**eFigure 6 in Supplemental 1**), with the strongest correlation being between IFN-γ response and neutralization capacity. This suggests that vaccine-induced immune protection relies on a synchronized adaptive immune response.^31,32^ To better depict the global immune response of each subject, we calculated an immunity score based on the presence or absence of RBD-binding antibodies, SARS-CoV-2 neutralizing antibodies and CMI response for each participant. We showed that none of the naïve individuals had a global immunity score of three after one dose of vaccine **(Figure 1B)**. In contrast, 84% of the previously infected individuals reached the maximal immunity score, the remainder showing an incomplete response and 6% demonstrating nearly no response **(Figure 1B)**.

### Symptomatology at the time of infection impacts the immune response to one vaccine dose

As 20% of recovered participants had an incomplete immune response after one dose of vaccine, we questioned if the presence of symptoms at the time of infection influenced the immune response to vaccination. Compared to symptomatic subjects, individuals who remained asymptomatic during their initial infection exhibited reduced anti-RBD IgG levels (7.3 ± 0.3 vs. 4.4 ± 0.5, P<.001), neutralizing antibody titers (592.5 ± 123.0 vs. 333.2 ± 119.1, P=.007) and IFN-γ secreting cells (344.9 ± 56.69 vs. 152.9 ± 35.1 SFU/10^6^ PBMCs, P=.02) (**Figure 2A)**. Importantly, asymptomatic individuals were less likely to neutralize SARS-CoV-2 after one dose of vaccine compared to symptomatic subjects (68% vs. 97%, P=.007). Furthermore, 92% of symptomatic individuals showed a global immune response with an immunity score of three after their first dose of vaccine compared to 69% of asymptomatic individuals **(Figure 2B)**.

**Figure 2:**
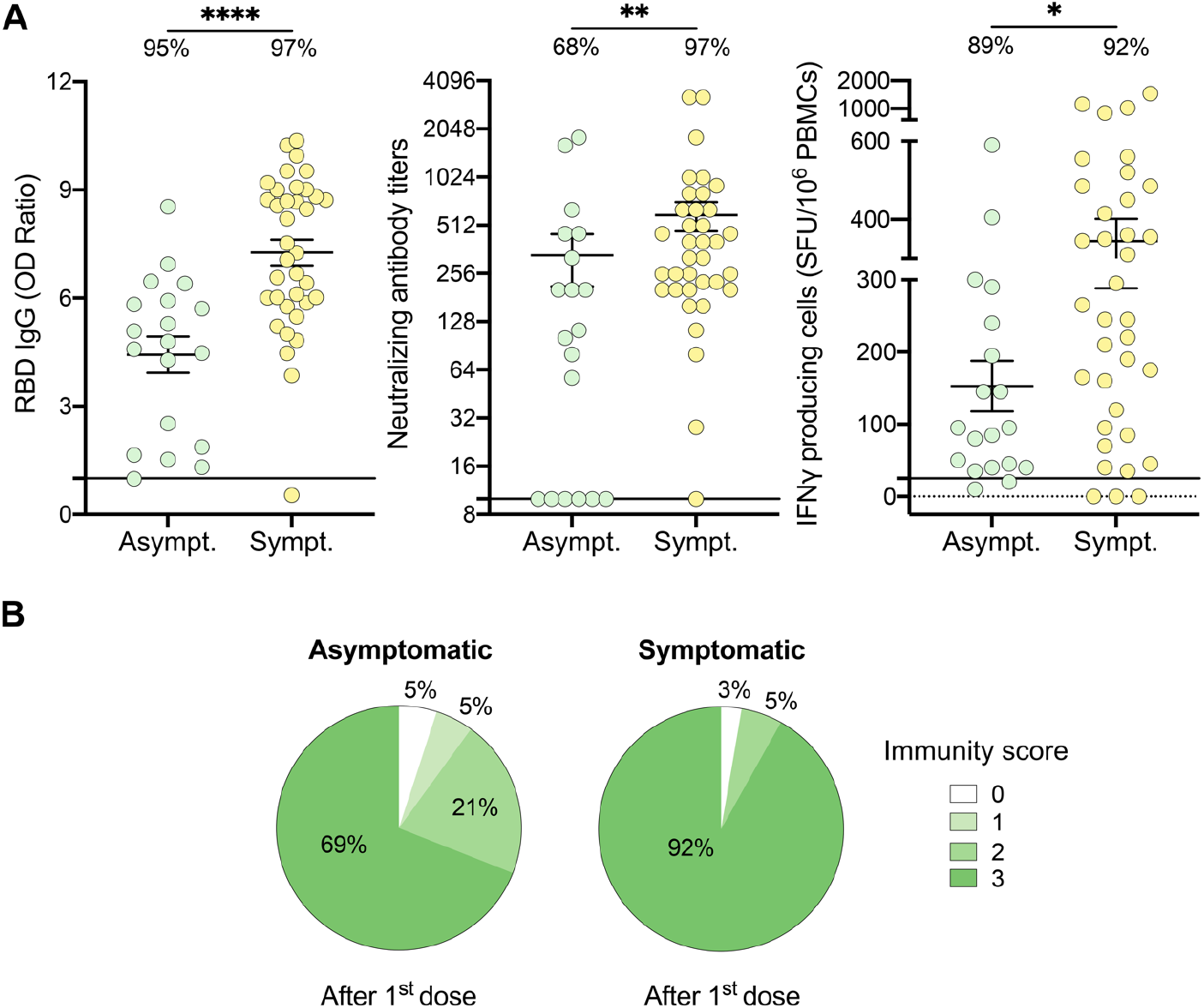
HCWs who recovered from an asymptomatic SARS-CoV-2 infection demonstrate partial immune response after one dose of vaccine. (**A**) SARS-CoV-2 Spike RBD–specific binding IgG levels assessed by ELISA (left panel), SARS-CoV-2 neutralizing antibody titers (middle panel), IFN-γ secreting cells per million in response to SARS-CoV-2 Spike glycoprotein peptides (right panel) assessed by ELISpot for asymptomatic (Asympt.) (n=19) and symptomatic (Sympt.) (n=36) recovered HCWs after one dose of vaccine. The black line indicates the positive threshold value, and the percentage of HCWs with responses above positive threshold value is indicated for each assay. Error bars indicate mean ± SEM. Statistical significance was assessed by Wilcoxon-Mann-Whitney tests. *P <.05; **P <.01; ***P <.001; ****P <.0001. (**B**) Pie charts of calculated immunity score for asymptomatic (left panel) and symptomatic (right panel) recovered HCWs after one dose of vaccine.

### Serology status six months after infection influences the strength of immune responses following one vaccine dose

In our RECOVER cohort, 24.6% (n = 140) of recovered HCWs were seronegative at enrollment in the study.^23^ We thus questioned if serostatus at enrollment, 3.0 ± 1.5 months prior to vaccination, could predict the immune response to the vaccine. Compared to seropositive individuals, seronegative subjects presented reduced levels of IgG (7.4 ± 0.3 vs. 4,8 ± 0.5, P<.001), neutralizing antibodies (739.9 ± 144.8 vs. 196.8 ± 47.6, P<.001), and IFN-γ producing cells (397.3 ± 61.5 vs. 125.2 ± 27.1, P<.001) after one dose of vaccine (**Figure 3A**). Importantly, subjects who did not mount a positive humoral and/or cellular response after one dose of vaccine were all seronegative before vaccination. One dose of vaccine was only sufficient to boost immunity to SARS-CoV-2 in 63% of seronegative participants **(Figure 3B)**. The others showed reduced capability to mount adequate neutralization of the virus and/or cell-mediated immune response, and two subjects remained nonresponsive.

**Figure 3:**
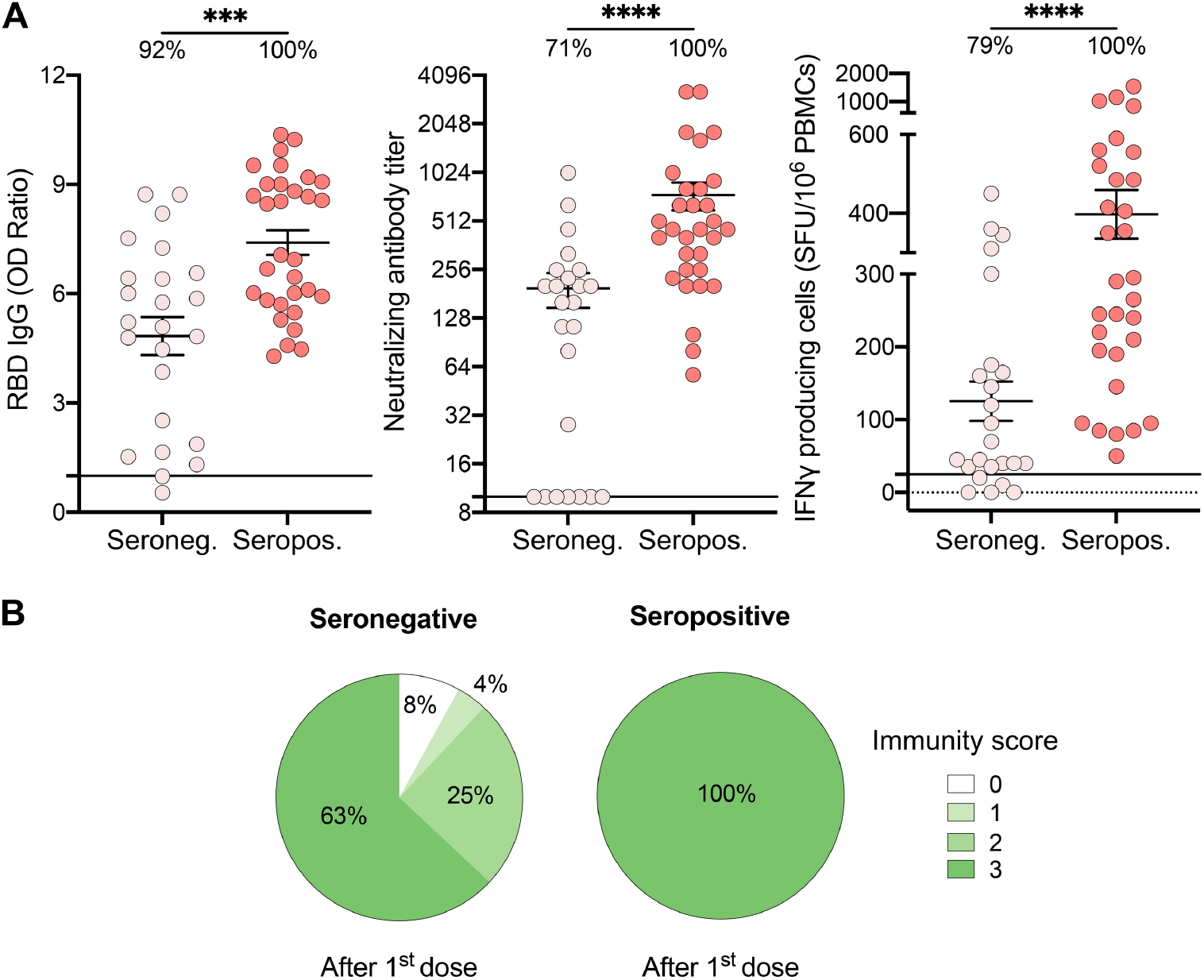
Recovered HCWs who were seronegative prior to vaccination showed suboptimal immune responses after one vaccine dose. (**A**) SARS-CoV-2 Spike RBD–specific binding IgG levels assessed by ELISA (left panel), SARS-CoV-2 neutralizing antibody titers (middle panel), IFN-γ secreting cells per million in response to SARS-CoV-2 Spike glycoprotein peptides (right panel) assessed by ELISpot for seronegative (Seroneg.) (n=24) and seropositive (Seropos.) (n=31) recovered HCWs after one dose of vaccine. The black line indicates the positive threshold value, and the percentage of HCWs with responses above positive threshold value is indicated for each condition. Statistical significance was assessed by Wilcoxon-Mann-Whitney tests. *P <.05; **P <.01; ***P <.001; ****P <.0001. (**B**) Pie charts of calculated immunity score for seronegative (left panel) and seropositive (right panel) recovered HCWs after one dose of vaccine.

### Combined effect of symptomatology and serostatus on the immunogenicity of the vaccine

Most previously infected HCWs who did not develop an immunity score of three after the first dose of vaccine had an asymptomatic SARS-CoV-2 infection (6 out of 9; **Figure 2B**). To better depict the impact of seropositivity and symptomatology on the immune response to one dose of vaccine, we compared levels of induced immunity within each subgroup. Symptomatic individuals who remained seropositive following natural infection developed the most vigorous humoral and cellular immune response to one dose of vaccine **(Figure 4A-B)**. Furthermore, their frequency of SARS-CoV-2 specific CD4^+^ T cells, as assessed by the expression of activation markers, was the highest (1.0 ± 0.2 vs. 0.3 ± 0.1 for naïve subjects, P=.006, **Figure 4C)**. In contrast, asymptomatic individuals who were seronegative at enrollment had significantly lower anti-RBD IgG levels (2.9 ± 0.6 vs. 8.2 ± 0.4, P<.001), neutralizing antibody titers (94.7 ± 50.0 vs. 831.4 ± 190.7, P<.001) as well as IFN-γ secreting cells (81.1 ± 30.7 vs. 482.9 ± 81.3 SFU/10^6^ PBMCs, P<.001) following one dose of the vaccine, compared to seropositive symptomatic subjects. Notably, this subgroup of asymptomatic seronegative individuals was the only group to demonstrate an immune response following one dose of vaccine that was similar to naïve individuals. In fact, prior to vaccination, these individuals lacked immunological markers of previous antigen encounter, as did SARS-CoV-2 naïve subjects (**eFigure 7 in Supplemental 1**).

**Figure 4:**
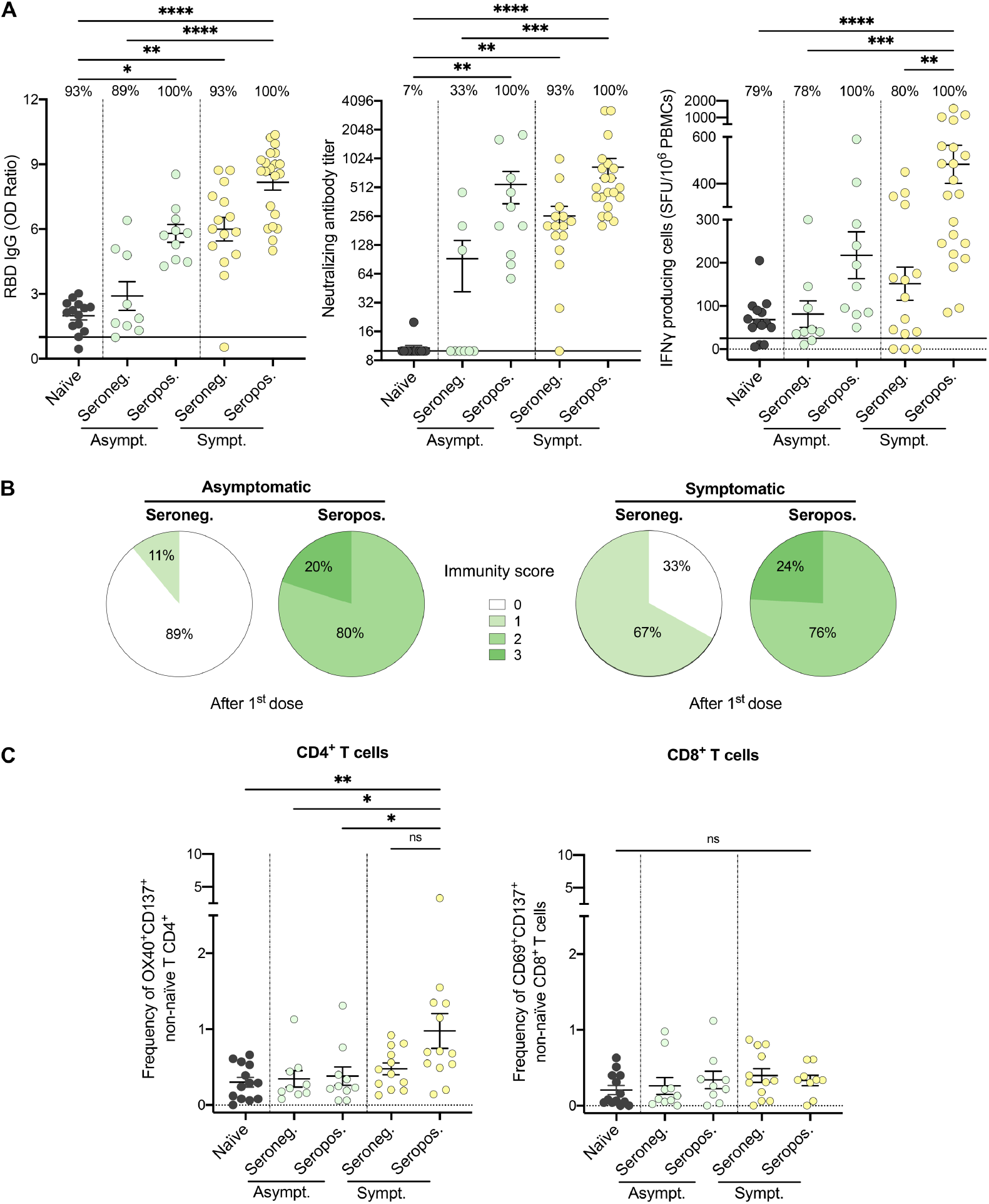
Both symptomatology during infection and serostatus prior to vaccination influence the immunogenicity of the first vaccine dose. (**A**) SARS-CoV-2 Spike RBD–specific binding IgG levels assessed by ELISA (left panel), SARS-CoV-2 neutralizing antibody titers (middle panel), and IFN-γ secreting cells per million in response to SARS-CoV-2 Spike glycoprotein peptides assessed by ELISpot (right panel) for naïve (black circles), seronegative (Seroneg.) (n=24) and seropositive (Seropos.) (n=31) recovered HCWs after one dose of vaccine. Asymptomatic recovered HCWs (Asympt.) are represented with green circles (n=19) and symptomatic HCWs (Sympt.) with yellow circles (n=36). The black line indicates the positive threshold value, and the percentage of HCWs with responses above positive threshold value is indicated for each assay. (**B**) Pie charts of calculated immunity score for asymptomatic (left panel) and symptomatic (right panel) recovered HCWs after one dose of vaccine. (**C**) Frequency of SARS-CoV-2−Spike-specific T cells measured as percentage of OX40^+^CD137^+^ non-naïve CD4^+^ (left panel) and CD69^+^CD137^+^ non-naïve CD8^+^ T cells (right panel) after stimulation of PBMCs with CD4-S mega pool of peptides from the Spike glycoprotein for naïve (n=13), asymptomatic seronegative (n=9) or seropositive (n=10) and symptomatic seronegative (n=12) or seropositive (n=13) recovered HCWs after one dose of vaccine. Error bars indicate mean ± SEM. Statistical significance was assessed by Kruskal-Wallis (**A-C**). *P <.05; **P <.01; ***P <.001; ****P <.0001.

### A two-dose primary series of mRNA vaccine is required in previously infected individuals who did not experience symptoms

Among HCWs who recovered from SARS-CoV-2 infection, 16 % did not demonstrate a well-coordinated humoral and cellular immune response after one vaccine dose **(Figure 1B)**, with asymptomatic, seronegative subjects being the least responsive. Therefore, we assessed humoral and cellular responses after the second dose of vaccine. Strikingly, all subjects displayed a strong humoral and cellular immune response after two doses of vaccine, including naïve and asymptomatic participants, as demonstrated by elevated anti-RBD IgG levels, neutralizing capability, as well as IFN-γ secreting cells (**Figure 5A** and **eFigure 8 in Supplemental 1**). After two doses of vaccine, naïve individuals still showed slightly lower anti-RBD IgG levels compared to both asymptomatic and symptomatic recovered individuals (**Figure 5B**). Cellular responses remained lower in asymptomatic HCWs compared to their symptomatic counterparts (**Figure 5B**). Our results collectively demonstrate that the severity of initial infection leads to the development of a more robust and persistent immune memory response after a vaccination challenge. In contrast, individuals that did not experience symptoms during infection with SARS-CoV-2 are less likely to develop long-term humoral and cellular immunity.

**Figure 5:**
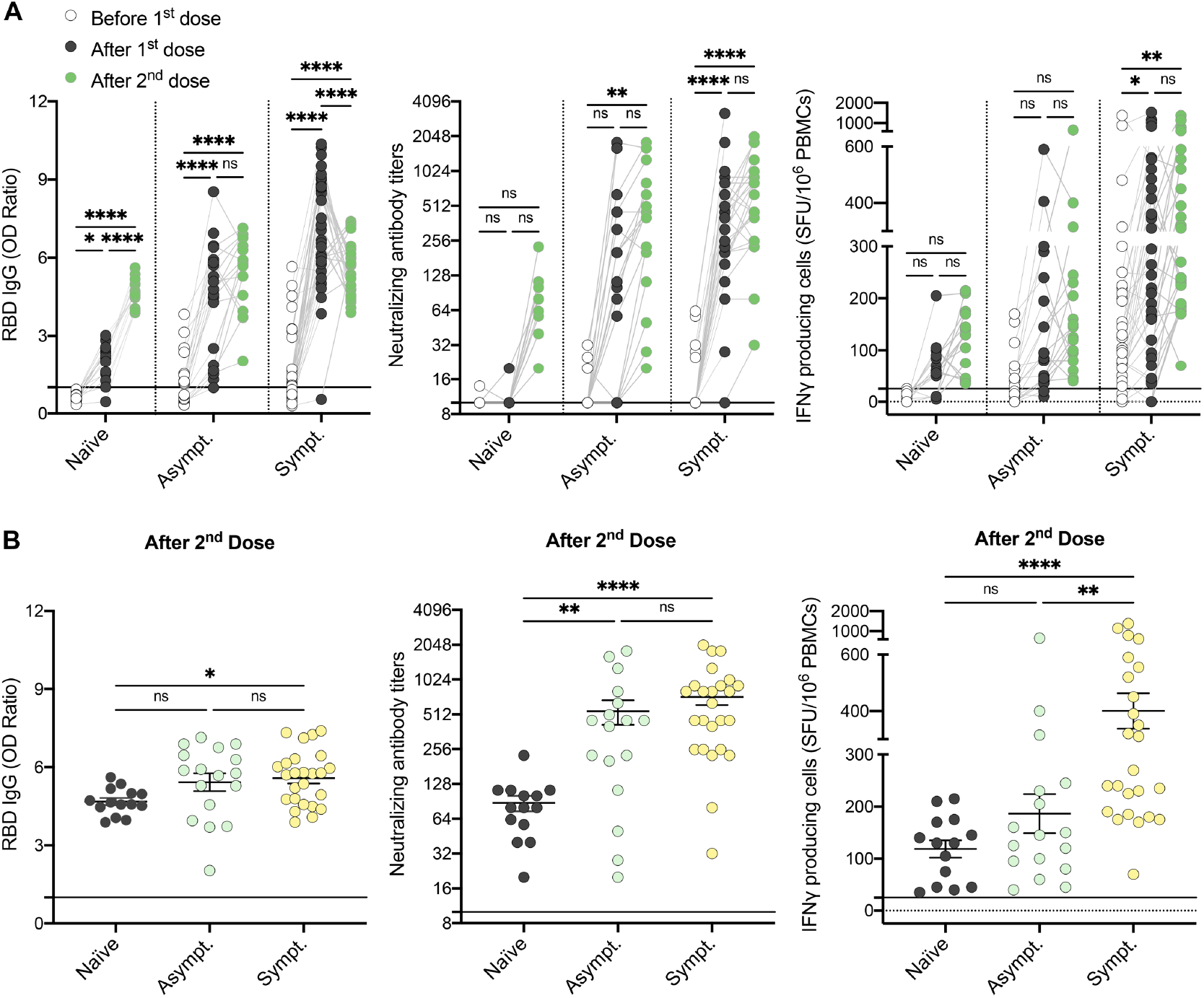
Two doses of vaccine are required for individuals who recovered from an asymptomatic SARS-CoV-2 infection to generate a global and coordinated adaptive immune response. (**A**) SARS-CoV-2 Spike RBD–specific binding IgG levels assessed by ELISA (left panel), SARS-CoV-2 neutralizing antibody titers (middle panel), IFN-γ secreting cells per million in response to SARS-CoV-2 Spike glycoprotein peptides (right panel) assessed by ELISpot for naïve (n=14), asymptomatic (n=17) and symptomatic recovered HCWs (n=25) before (white circles), after one dose (black circles) and after two doses of vaccine (green circles). (**B**) SARS-CoV-2 Spike RBD–specific binding IgG levels assessed by ELISA (left panel), SARS-CoV-2 neutralizing antibody titers (middle panel), and IFN-γ secreting cells per million in response to SARS-CoV-2 Spike glycoprotein peptides assessed by ELISpot (right panel) for naïve (black circles), asymptomatic (n=17) and symptomatic (n=25) recovered HCWs after two doses of vaccine. The black line indicates the positive threshold value. Error bars indicate mean ± SEM. Kruskal-Wallis tests assessed statistical significance. *P <.05; **P <.01; ***P <.001; ****P <.0001.

## DISCUSSION

The ultimate goal of vaccination is to generate broad and long-lasting immune responses that will protect the host from severe clinical outcomes if infected. The most appropriate vaccination regimen to be used in previously infected individuals remains undefined, leading to discrepancies in public health recommendations worldwide. Based on prior studies suggesting that previously infected individuals did not benefit from their second dose of mRNA vaccine,^18,22^ many countries have moved forward with the recommendation that a single dose of mRNA vaccine was sufficient for individuals who were previously infected.^9^ In this report, we demonstrate that one dose of vaccine is sufficient to induce sustained humoral and cellular immune responses in 92% of previously infected individuals who experienced symptoms during their infection. In contrast, only 69% in individuals who remained asymptomatic during their primary infection mounted robust immune responses to one dose of vaccine. In fact, infected individuals who remained asymptomatic and were seronegative at enrollment in our study harbored an immune response comparable to naïve subjects before and after vaccination. This is true even when using an extended vaccination regimens shown to favor better immune memory,^33^ as our cohort was vaccinated with a 16-week interval between each vaccine dose.

Our results differ from one American HCWs cohort that concluded that humoral immune responses shortly after one mRNA vaccine dose were comparable in individuals that recovered from symptomatic and asymptomatic infections.^34^ However, HCWs from that cohort were all seropositive at the time of vaccination. Here, we show that previously infected but asymptomatic individuals that harbour a negative serostatus at vaccination mount reduced serum neutralization capabilities and cell-mediated memory responses following one vaccine dose. Our report supports the requirement for a complete vaccination regimen for adequate protection of individuals who do not experience symptoms during their SARS-CoV-2 infection.

Knowledge gained from this study is particularly important in the current context of booster vaccination and protection against recent variants-of-concerns (VOCs). Waning immunity has been reported six months after a two-dose primary series in naïve individuals.^35,36^ In these naïve individuals, three doses of vaccine increase protection against symptomatic and severe B.1.617.2 (Delta) and B.1.1.529 (Omicron) infections.^37-42^ Notably, despite strengthened immune responses following the booster dose, the capacity to neutralize the B.1.1.529 variant appears weaker.^43-46^ A German study reported that neutralizing antibodies against the Omicron variant were highest in individuals who had encountered the antigen three times, having received two doses of vaccine and having been infected with SARS-CoV-2 prior to or subsequent to vaccination (hybrid immunity) or having received three doses of mRNA vaccine for uninfected individuals.^47^ Based on our current report, previously infected individuals who did not develop symptoms should be prioritized in getting a booster shot, as should be previously uninfected ones, as we demonstrate that a fraction of these individuals harbor immune responses to vaccination comparable to naïve individuals. Further studies will be required to evaluate the vaccine requirements for boosters in recently infected Omicron cases, as people infected with this variant are more likely to be asymptomatic.^48^

### Limitations

Results of this study were obtained from a cohort of primarily middle-aged Caucasian females infected with SARS-CoV-2 eight months before vaccination. It remains to be evaluated if a shorter time between infection and vaccination will lead to similar results. Our study is also limited by a relatively small sample size of participants who did not experience symptoms during their initial infection. It will be essential to validate our findings in a larger population of subjects, as immune responses to SARS-CoV-2 are known to be highly heterogeneous. Furthermore, the protection induced by booster doses against VOCs in previously infected individuals remains to be evaluated. This study did not assess the impact of other approved adenovirus or mRNA vaccines against SARS-CoV-2^49,50^, as primary series or boosters, as well as natural infection with emerging SARS-CoV-2 VOCs, as most of the cohort was infected at the beginning of 2020.^51^ Finally, the immunity score may underestimate the quality of the immune memory generated in recovered individuals since it is limited to three of the most standardized assays to interrogate immune response to SARS-CoV-2. Detailed functional, transcriptomic and repertoire analyses will be essential to fully grasp the immune memory differences between individuals who experienced symptoms or not following infection.

## Conclusions

This report reveals the critical importance of the impact of an individual’s symptomatology at the time of infection and serostatus at the time of vaccination on the capacity to mount adequate immune protection, including cell-mediated responses. In contrast to individuals who were symptomatic during infection, we demonstrated that a third of the individuals who recovered from an asymptomatic infection responded like naïve individuals. Two doses of vaccine should be offered to previously infected but asymptomatic individuals to elicit a global and sustained adaptive immune response following mRNA vaccination.

## Data Availability

All data produced in the present study are available upon reasonable request to the authors

## Acknowledgment Section

We want to thank Valérie Villeneuve, Jocelyne Ayotte, and the Rare Pediatric Disease (RaPID) Biobank personnel at the CHU Sainte-Justine for their technical help and expertise. Special thanks to Fazia Tadount and the team at the Vaccine Study Center for coordinating the RECOVER studies, and to Zineb Laghdir for administrative support for the RECOVER studies. We are grateful to the nurses and research coordinators at each University Hospital for their involvement in this study and the healthcare workers who agreed to participate in this research. We also thank Alessandro Sette and Daniela Weiskopf for providing the mega pool of peptides utilized in the AIM assays.

## Author Contributions

HD, CQ conceptualized the project, designed the study and experiments, and obtained funding

SN, BB, JC, HR, MEH performed the experiments

SN, BB, HD, MB analyzed the data

SN, BB, HD interpreted the results and prepared the figures

KA, DM performed the clinical follow-up of the study

SN, BB, HD wrote the manuscript

CQ, VG, MPC, PS, GDS, JC, YL, MB, GB edited the manuscript and provided critical input

