## Supplemental Tables and Figures for "Immunogenicity of Pfizer-BioNTech COVID-19 mRNA Primary Vaccination Series in Recovered Individuals Depends on Symptoms at Initial Infection"

**SUPPLEMENTAL FIGURES AND FIGURE LEGENDS**

3 Supplemental Tables

**Supplemental Table 1:** Characteristics of naïve and recovered HCWs included in assays.

**Supplemental Table 2:** Detailed time points for assessment of naïve individuals and HCWs who recovered from previous SARS-CoV-2 infection and were vaccinated with BNT162b2 mRNA vaccine.

**Supplemental Table 3:** Antibodies used for flow cytometry analysis.

9 Supplemental Figures

**eFigure 1:** Timeline of blood collection for humoral and cellular analyses of vaccinated HCWs who recovered from SARS-CoV-2 infection or individuals naïve to SARS-CoV-2 before vaccination.

**eFigure 2:** Gating strategy for AIM assays and representative dot plots.

**eFigure 3:** The strength of the humoral response in recovered HCWs from SARS-CoV-2 infection is correlated with symptomatology during infection.

**eFigure 4:** Recovered HCWs from SARS-CoV-2 infection maintain strong and stable SARS-CoV-2-specific cellular memory up to 11 months after infection.

**eFigure 5:** Recovered HCWs develop much stronger SARS-CoV-2-specific humoral and cellular immune responses than naïve individuals after one dose of vaccine.

**eFigure 6:** Humoral and cellular responses that developed after one dose of vaccine are correlated in HCWs who recovered from SARS-CoV-2 infection.

**eFigure 7:** Both humoral and cellular natural responses after SARS-CoV-2 infection are significantly impaired in seronegative individuals.

**eFigure 8:** Previously infected individuals develop stronger humoral and cellular responses to mRNA vaccination than naïve subjects.

**SUPPLEMENTAL TABLES**

**Supplemental Table 1: Characteristics of naïve and recovered HCWs included in assays.**

|  | **Number (%) of naïve individuals (n=14)** | **Number (%) of recovered HCWs (n=55)** |
| --- | --- | --- |
| **Demographics** |  |  |
| **Sex** |  |  |
| Male | 5 (36) | 20 (36) |
| Female | 9 (64) | 35 (64) |
| **Age (y)** |  |  |
| Mean ± SD | 43 ± 9 | 41.8 ± 12.3 |
| Median [Min, Max] | 41 [27, 57] | 44.6 [19.4, 67.1] |
| **Ethnicity** |  |  |
| Caucasian | 13 (93) | 45 (82) |
| Middle Eastern | 0 (0) | 1 (2) |
| Latin American | 0 (0) | 2 (4) |
| Asian | 1 (7) | 4 (7) |
| Black or African American | 0 (0) | 3 (5) |
| **Symptomatology at initial infection** |  |  |
| Asymptomatic / WHO Score of 1 | n/a | 19 (35) |
| Symptomatic / WHO Score of 2-3 | n/a | 30 (54) |
| Symptomatic / WHO Score of 4-5 | n/a | 6 (11) |
| **Time since initial infection at enrollment (m)** |  |  |
| Mean ± SD | n/a | 5.3 ± 1.9 |
| Median [Min, Max] | n/a | 5.3 [0.7, 9.3] |
| **Serology Status at Enrollment** |  |  |
| **Seronegative** | n/a | 24 (44) |
| **Seropositive** | n/a | 31 (56) |
| SD: Standard Deviation. |  |  |
| y: year, m: months. |  |  |

**Supplemental Table 2: Detailed time points for assessment of naïve individuals and HCWs who recovered from previous SARS-CoV-2 infection and were vaccinated with BNT162b2 mRNA vaccine.**

|  | **Number (%) of naive individuals (n=14)** | **Number (%) of recovered HCWs (n=55)** |
| --- | --- | --- |
| **Vaccine received** |  |  |
| Pfizer BioNTech | 14 (100) | 55 (100) |
| **Time since initial infection at pre-vaccination analysis (m)** |  |  |
| Mean ± SD | n/a | 7.0 ± 2.2 |
| Median [Min, Max] | n/a | 7.1 [0.7, 13.1] |
| **Time since initial infection at first vaccine dose (m)** |  |  |
| Mean ± SD | n/a | 8.4 ± 2.3 |
| Median [Min, Max] | n/a | 8.6 [0.8, 13.1] |
| **Time since first vaccine dose at analysis (d)** |  |  |
| Mean ± SD | 29.1 ± 2.1 | 45.1 ± 25.1 |
| Median [Min, Max] | 28 [27, 33] | 35 [10, 93] |
| **Time since initial infection at second vaccine dose (m)** |  |  |
| Mean ± SD | n/a | 12.4 ± 2.5 |
| Median [Min, Max] | n/a | 12.6 [5.0, 16.4] |
| **Time since second dose at analysis (d)** |  |  |
| Mean ± SD | 35.2 ± 4.9 | 56.8 ± 30.4 |
| Median [Min, Max] | 34 [26.45] | 52 [13, 121] |
| **Time between first and second dose  vaccination (w)** |  |  |
| Mean ± SD | 9.5 ± 0.8 | 16.2 ± 5.6 |
| Median [Min, Max] | 9.6 [8.0, 11.3] | 16.0 [3.0, 32.3] |
| SD: Standard Deviation. |  |  |
| w: weeks, m: months, d: days. |  |  |

**Supplemental Table 3: Antibodies used for flow cytometry analysis.**

| **Antibodies** | **Company** | **Clone** | **Cat#** |
| --- | --- | --- | --- |
| BUV395 Mouse Anti-Human CD3 | BD | SK7 | 564001 |
| V500 Mouse Anti-Human CD4 | BD | RPA-T4 | 560768 |
| AF700 Mouse Anti-Human CD8 | BD | RPA-T8 | 557945 |
| [BUV737 Mouse Anti-Human CCR7](https://www.bdbiosciences.com/us/reagents/research/antibodies-buffers/immunology-reagents/anti-human-antibodies/cell-surface-antigens/buv737-mouse-anti-human-ccr7-cd197-2-l1-a/p/749676) | BD | 2-L1-A | 749676 |
| eF450 Mouse Anti-Human CD45-RA | Invitrogen | HI100 | 48-0458-42 |
| APC Mouse Anti-Human CD137 | BioLegend | 4B4-1 | 309810 |
| PE-Cy7 Mouse Anti-Human OX40 | BioLegend | Ber-ACT35 | 350012 |
| BV785 Mouse Anti-Human CD25 | BioLegend | BC96 | 302638 |
| PE Mouse Anti-Human CD69 | BD | FN50 | 555531 |

**eFigure 1: Timeline of blood collection for humoral and cellular analyses of vaccinated HCWs who recovered from SARS-CoV-2 infection or individuals naïve to SARS-CoV-2 before vaccination.**

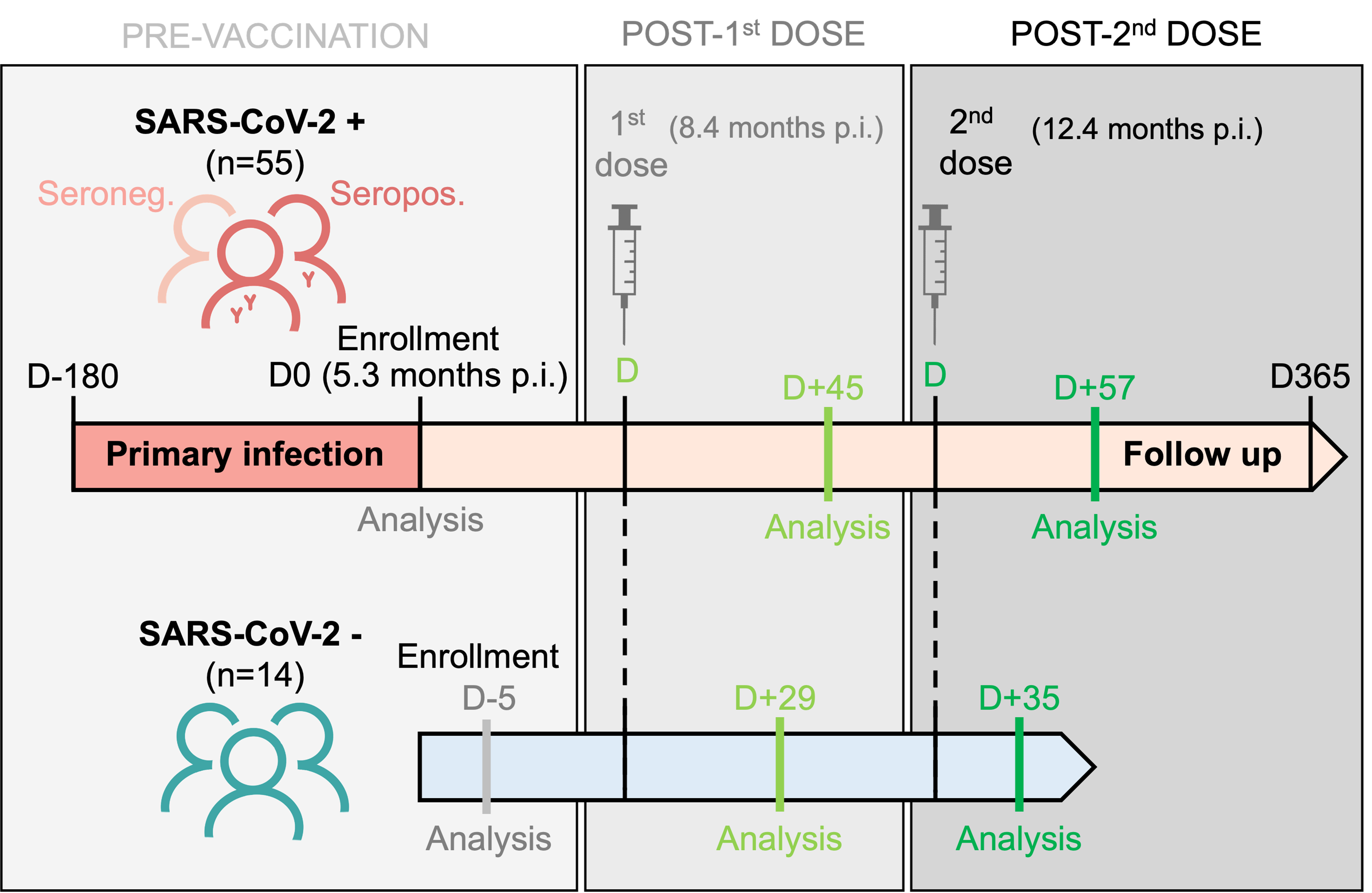

Blood samples for humoral and cell-mediated immunity were collected at enrollment (D0), around D+45 after the first dose and around D+57 after the second dose of vaccine for seronegative (Seroneg.) and seropositive (Seropos.) HCWs recovered from SARS-CoV-2 infection (SARS-CoV-2 +, n=55). For 14 naïve participants (SARS-CoV-2 -), blood samples were collected before their first dose of vaccine (D-5/D0), around D+29 after the first dose and around D+35 after the second dose.

**eFigure 2: Gating strategy for AIM assays and representative dot plots.**

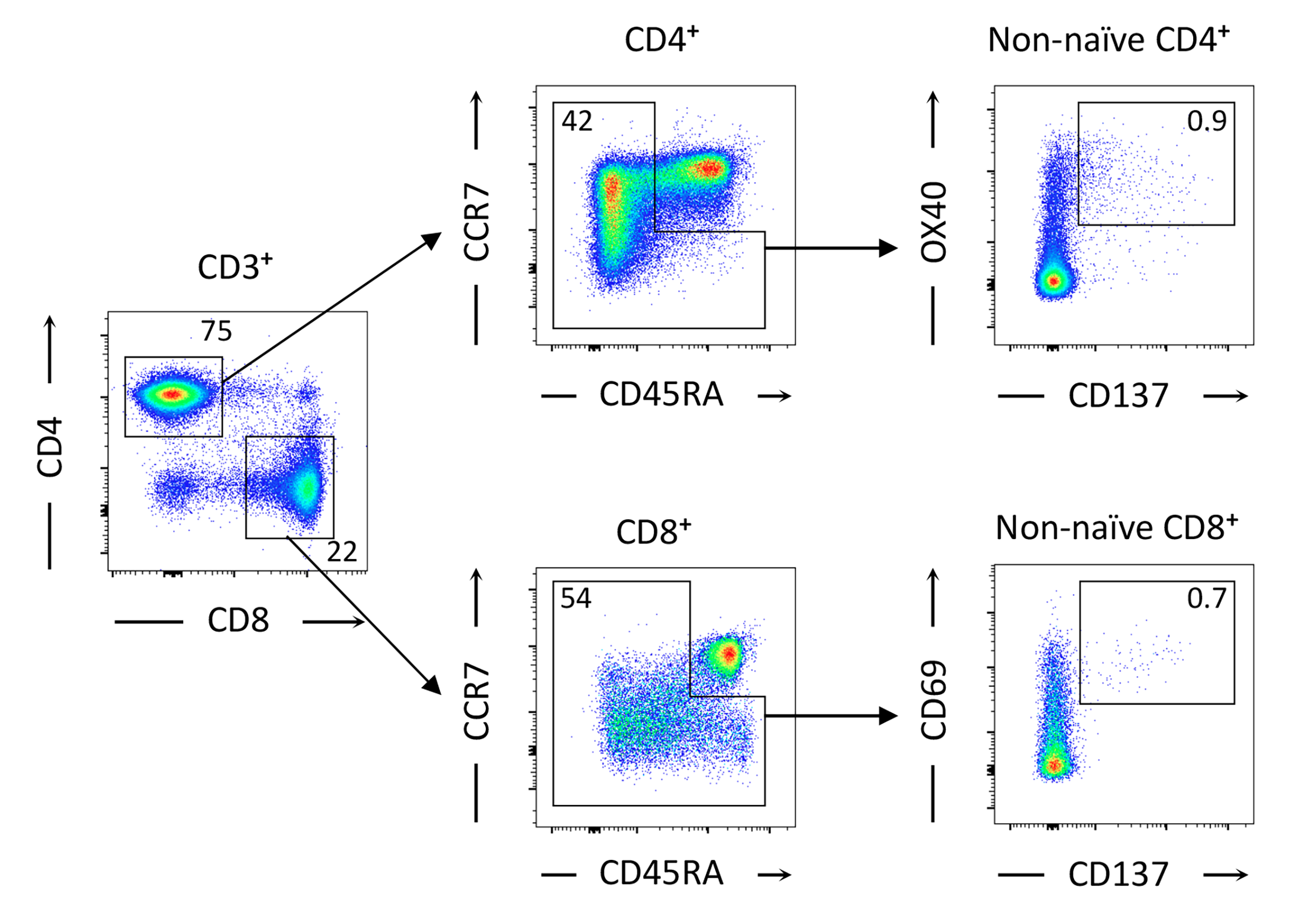

Gating used to identify SARS-CoV-2−specific CD4^+^ (OX40^+^CD137^+^) and CD8^+^ (CD69^+^CD137^+^) T cells in recovered HCWs from SARS-CoV-2 infection or naïve to the infection vaccinees after stimulation of PBMCs with CD4-S mega pool of peptides from the Spike glycoprotein. The percentage of events is indicated within AIM^+^ (double-positive) quadrant.

**eFigure 3: The strength of the humoral response in recovered HCWs from SARS-CoV-2 infection is correlated with symptomatology during infection.**

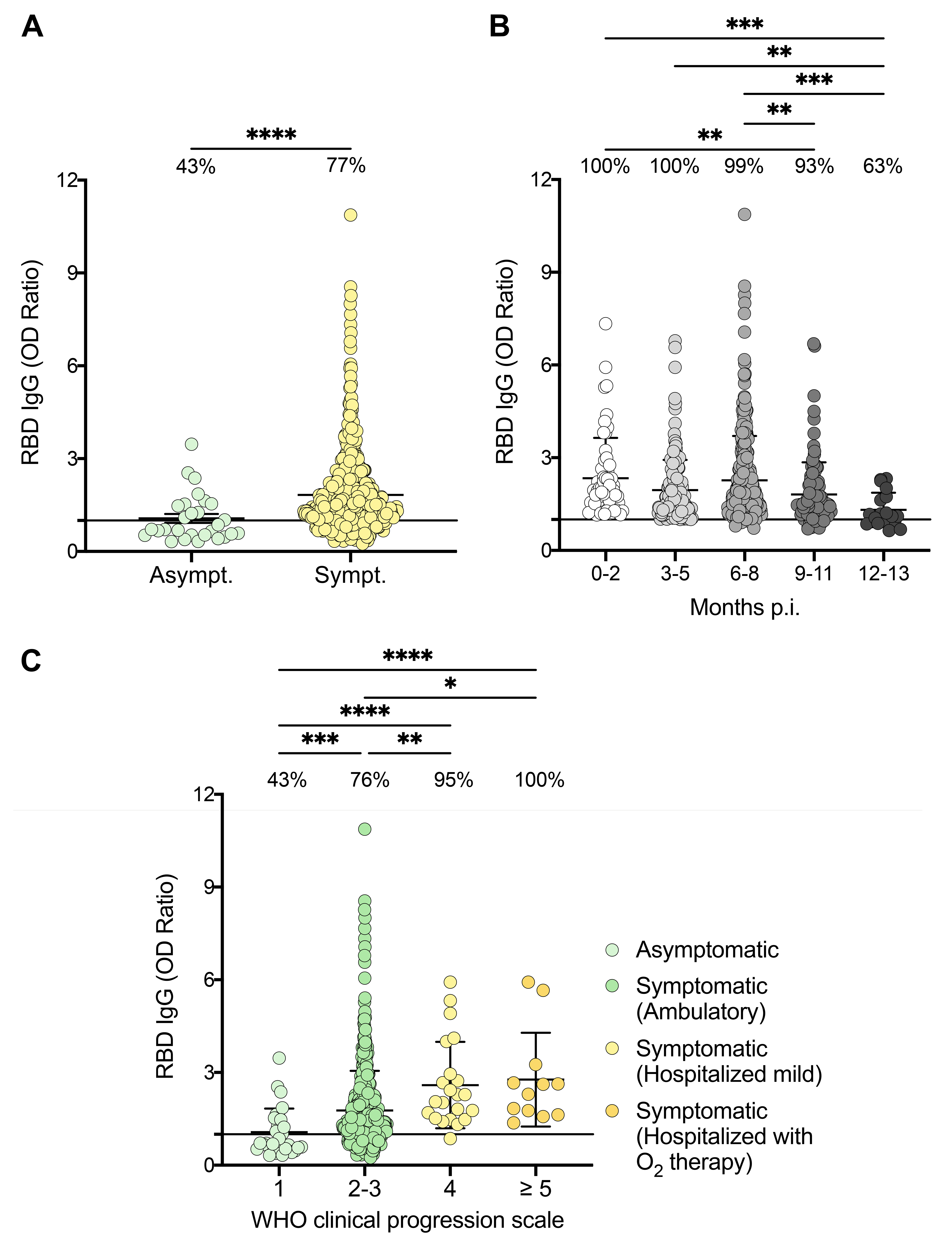

(**A**) SARS-CoV-2 Spike RBD–specific binding IgG levels assessed by ELISA for 569 recovered HCWs classified based on their symptomatology upon infection. (**B**) Kinetic of RBD-binding IgG levels over one-year post infection in participants who were seropositive at enrollment. (**C**) SARS-CoV-2 Spike RBD–specific binding IgG levels assessed by ELISA for 569 recovered HCWs classified according to WHO clinical progression scale. (**A**) Asymptomatic (Asympt.): n=28, Symptomatic (Sympt.): n=541. (**B**) 0-2 months: n=57, 3-5: n=175, 6-8: n=297, 9-11: n=108, 12-13: n=19. (**C**) WHO clinical progression scale 1: n=28, 2-3: n=507, 4: n=22, ≥5: n=12. The black line indicates the positive threshold value, and the percentage of seropositive participants is indicated for each subgroup. Error bars indicate mean ± SD. Statistical significance was assessed by Kruskal-Wallis (**A**) and Wilcoxon-Mann-Whitney (**B-C**) tests. *P <.05; **P <.01; ***P <.001; ****P <.0001.

**eFigure 4: Recovered HCWs from SARS-CoV-2 infection maintain strong and stable SARS-CoV-2-specific cellular memory up to 11 months after infection.**

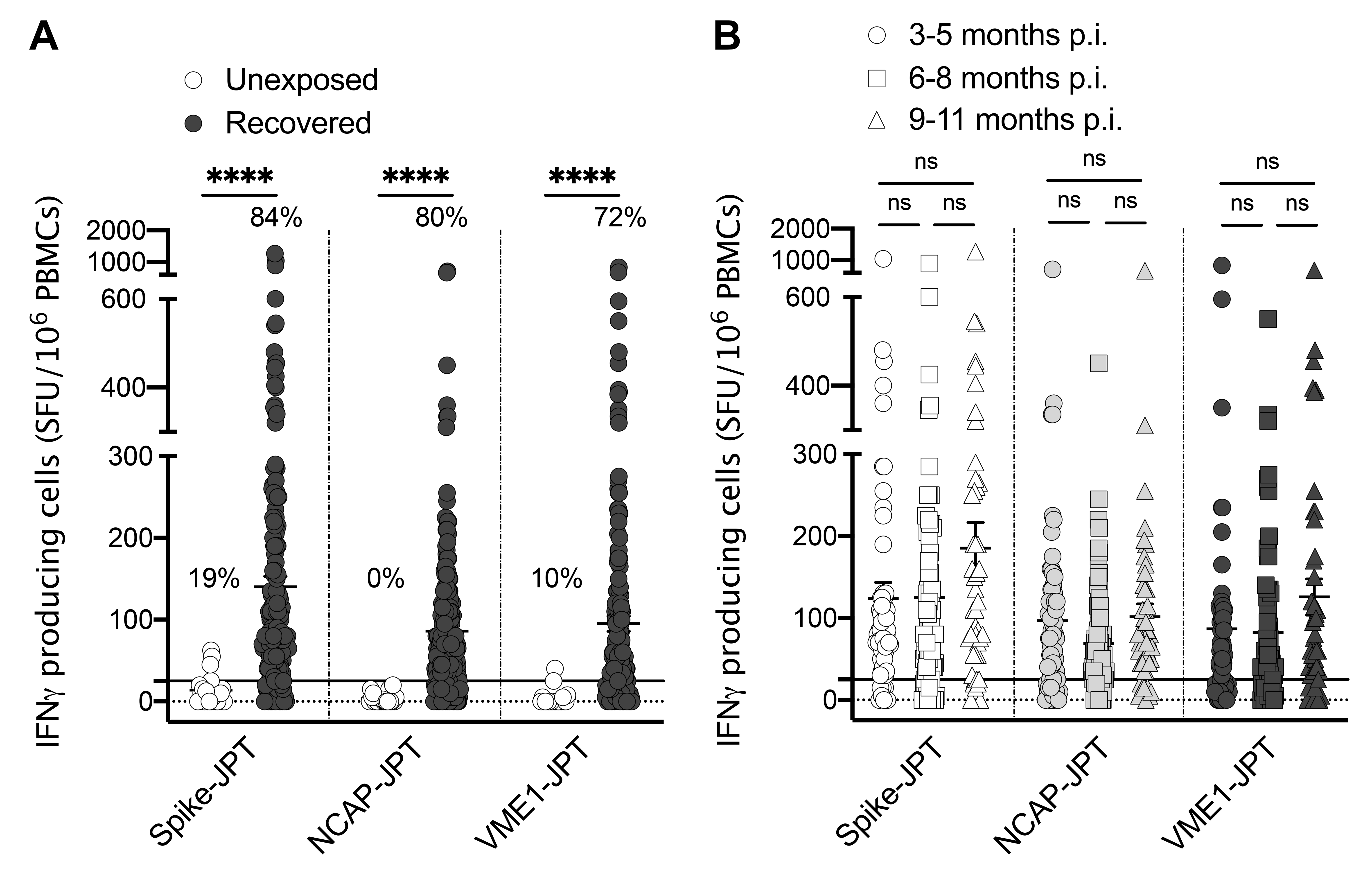

(**A-B**) IFN-γ producing cells per million in response to SARS-CoV-2 Spike glycoprotein, nucleocapsid protein (NCAP) and membrane protein (VME1) mega pools of peptides assessed by ELISpot for naïve (white circles) (n=21) and recovered individuals (black circles) (n=200) at enrollment (**A**) and kinetic of the response one-year post infection (3-5 months p.i.: n=63, 6-8 months p.i.: n=70, 9-11 months p.i.: n=48) (**B**). The black line indicates the positive threshold value, and the percentage of individuals with responses above positive threshold value is indicated for each subgroup and time point. Error bars indicate mean ± SD. Statistical significance was assessed by Wilcoxon-Mann-Whitney (**A**) and Kruskal-Wallis (**B**) tests. *P <.05; **P <.01; ***P <.001; ****P <.0001.

**eFigure 5: Recovered HCWs develop much stronger SARS-CoV-2-specific humoral and cellular immune responses than naïve individuals after one dose of vaccine.**

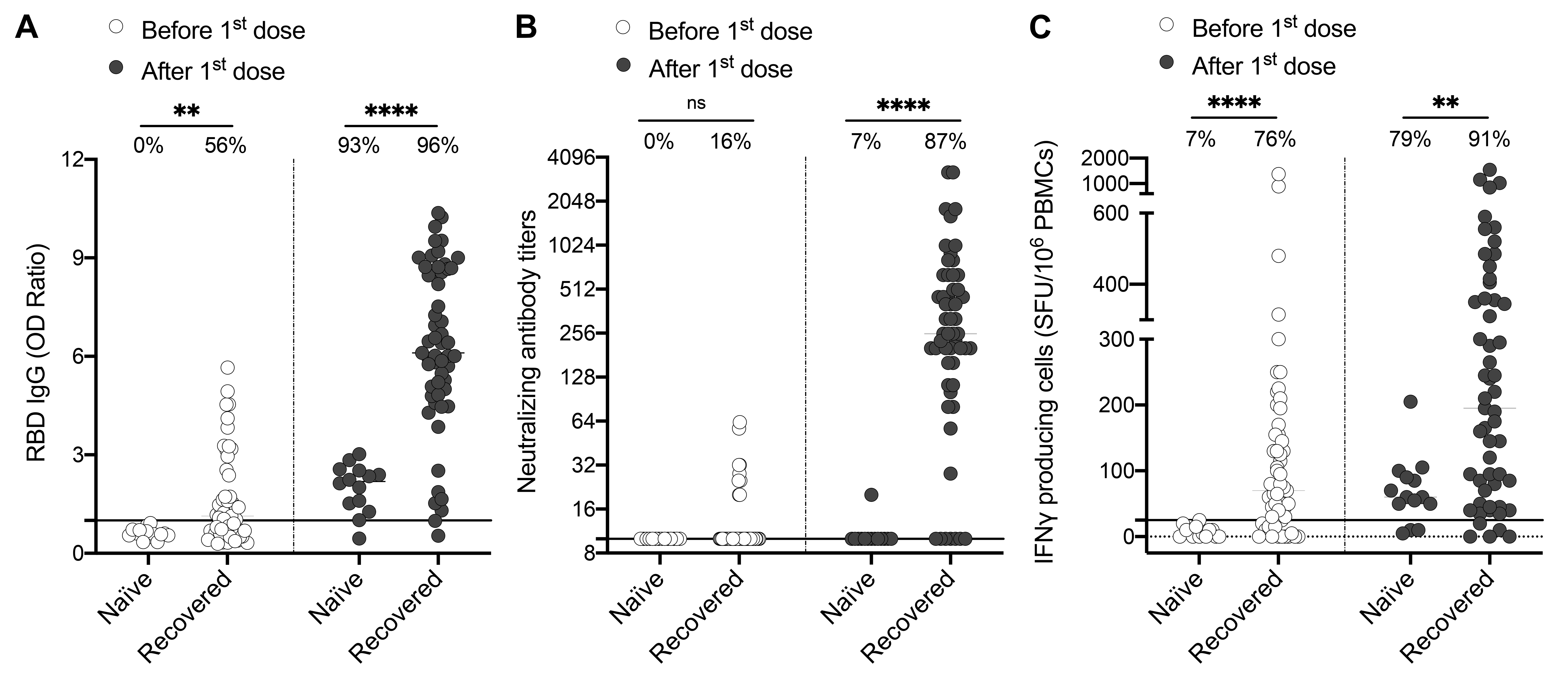

(**A-C**) SARS-CoV-2 Spike RBD–specific binding IgG levels assessed by ELISA (**A**), SARS-CoV-2 neutralizing antibody titers (**B**), IFN-γ producing cells per million in response to SARS-CoV-2 spike glycoprotein peptides stimulation (**C**) assessed by ELISpot for naïve (n=14) and recovered HCWs (n=55) before (white circles) and after one dose of vaccine (black circles). The black line indicates the positive threshold value, and the percentage of individuals with responses above positive threshold value is indicated for each subgroup and time point. Error bars indicate mean ± SEM. Statistical significance was assessed by Wilcoxon-Mann-Whitney tests. *P <.05; **P <.01; ***P <.001; ****P <.0001.

**eFigure 6: Humoral and cellular responses developed after one vaccine dose are correlated in HCWs who recovered from SARS-CoV-2 infection.**

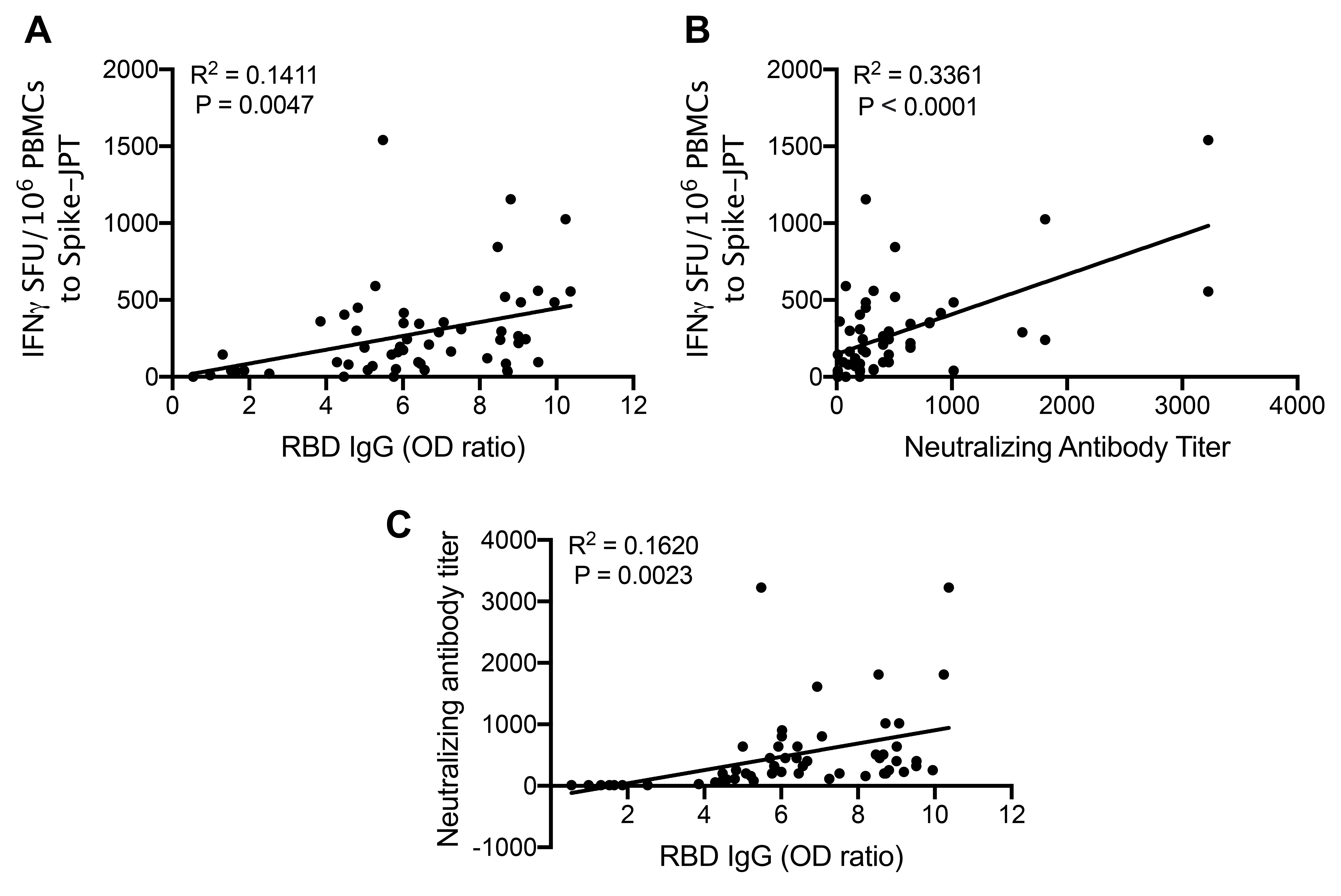

(**A-C**) Correlation between IFN-γ secreting cells per million in response to SARS-CoV-2 Spike glycoprotein peptides and SARS-CoV-2 Spike RBD–specific binding IgG levels (**A**), IFN-γ secreting cells per million in response to SARS-CoV-2 Spike glycoprotein peptides and SARS-CoV-2 neutralizing antibody titers (**B)**, SARS-CoV-2 neutralizing antibody titers and SARS-CoV-2 Spike RBD–specific binding IgG levels (**C**) in recovered HCWs from SARS-CoV-2 infection after one dose of vaccine (n=67). Correlation was assessed by Pearson correlation coefficient. Coefficient R^2^ and P-values are shown.

**eFigure 7: Both humoral and cellular natural responses after SARS-CoV-2 infection are significantly impaired in seronegative individuals.**

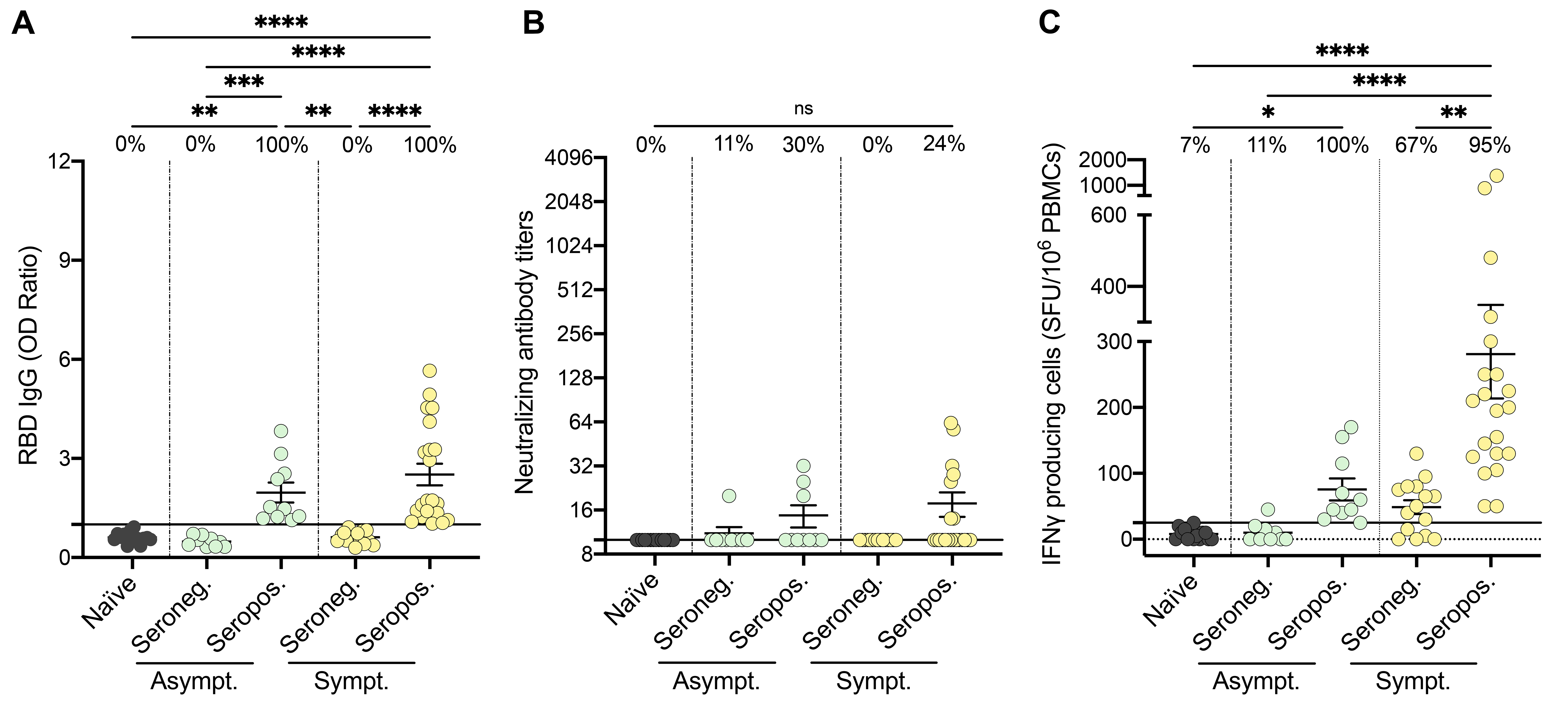

(**A-C**) SARS-CoV-2 Spike RBD–specific binding IgG levels assessed by ELISA (**A**), SARS-CoV-2 neutralizing antibody titers (**B**), IFN-γ secreting cells per million in response to SARS-CoV-2 Spike glycoprotein peptides stimulation (**C**) assessed by ELISpot for seronegative (Seroneg.) (n=18) and seropositive (Seropos.) (n=37) recovered HCWs after natural infection. Asymptomatic recovered HCWs (Asympt.) are represented with green circles (n=17) and symptomatic HCWs (Sympt.) with yellow circles (n=38). Naïve individuals are shown as negative control (black circles). The black line indicates the positive threshold value, and the percentage of HCWs with responses above positive threshold value is indicated for each subgroup. Error bars indicate mean ± SEM. Statistical significance was assessed by Kruskal-Wallis tests. *P <.05; **P <.01; ***P <.001; ****P <.0001.

**eFigure 8: Previously infected individuals develop stronger humoral and cellular responses to mRNA vaccination than naïve subjects.**

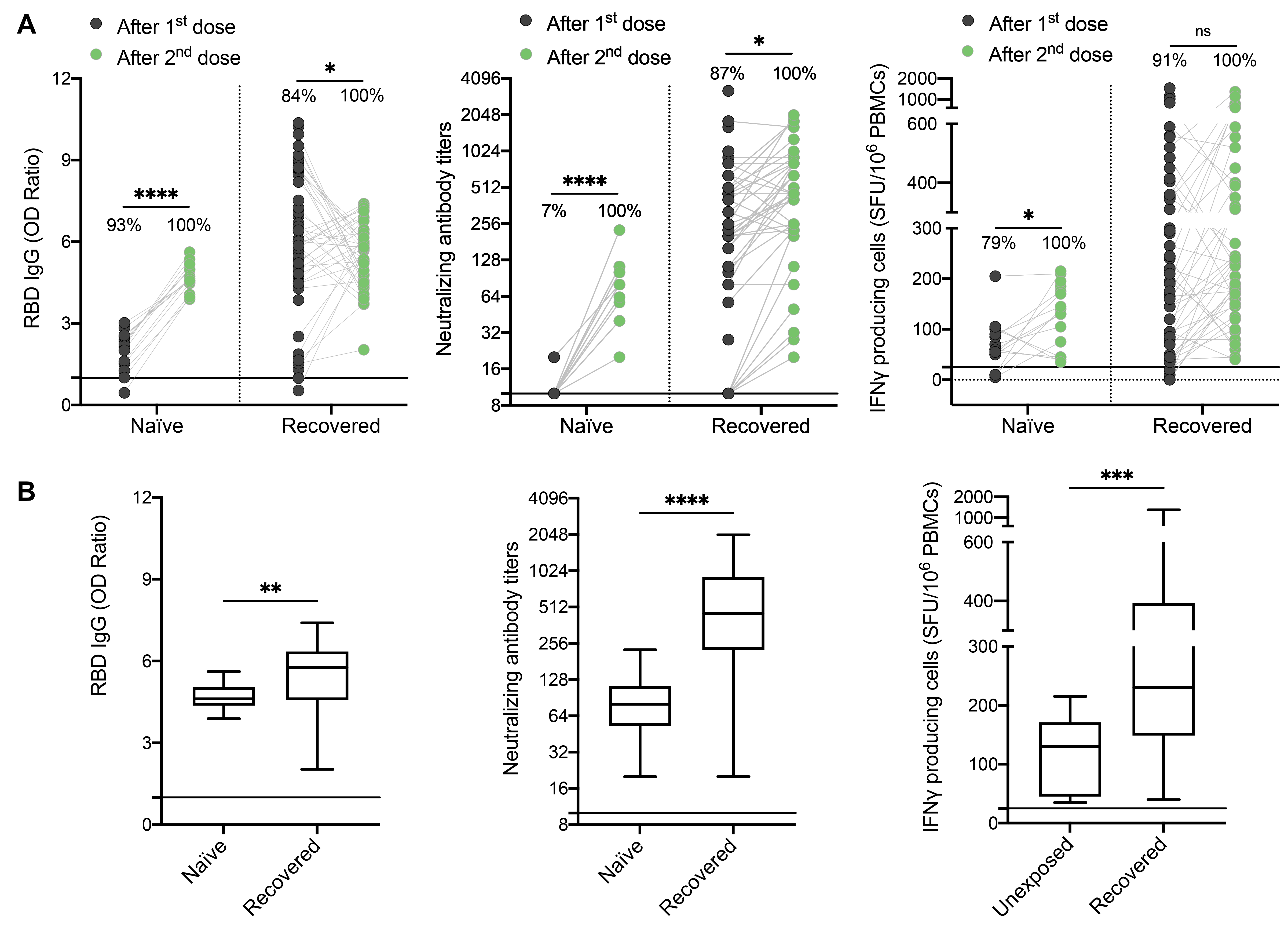

(**A**) SARS-CoV-2 Spike RBD–specific binding IgG levels assessed by ELISA (left panel), SARS-CoV-2 neutralizing antibody titers (middle panel), IFN-γ secreting cells per million in response to SARS-CoV-2 Spike glycoprotein peptides stimulation (right panel) assessed by ELISpot for naïve (n=14) and recovered individuals (n=55) after one dose (black circles) and two doses of vaccine (green circles). (**B**) SARS-CoV-2 Spike RBD–specific binding IgG levels assessed by ELISA (left panel), SARS-CoV-2 neutralizing antibody titers (middle panel), IFN-γ secreting cells per million in response to SARS-CoV-2 Spike glycoprotein peptides stimulation (right panel) assessed by ELISpot for naïve (n=14) and recovered individuals (n=42) after two doses of vaccine. The black line indicates the positive threshold value, and the percentage of individuals with responses above positive threshold value is indicated for each time point. Error bars indicate mean ± SEM. Statistical significance was assessed by Wilcoxon-Mann-Whitney tests. *P <.05; **P <.01; ***P <.001; ****P <.0001.
